# Genetic versus Modifiable Risk: Predicting Dementia, Cognition, and Brain Structure in the UK Biobank

**DOI:** 10.64898/2026.01.23.26344680

**Authors:** Yidan Zhang, Gamze Erzin, Colin Rosenau, Sebastian Köhler, Dennis van der Meer, Jurjen J. Luykx, David E. J. Linden, Bart P. F. Rutten, Sinan Guloksuz, Gabriëlla A. M. Blokland

## Abstract

**Introduction:** Both genetic and modifiable lifestyle factors contribute to dementia risk, yet their separate and joint predictive value for dementia and its intermediate markers (cognitive function and brain structure), is unclear. This study evaluated APOE ε4 status, AD polygenic risk (excluding APOE), and the updated LIBRA2 index (14 modifiable factors) in predicting dementia, cognition, and brain imaging biomarkers.

**Methods:** To examine associations of *APOE* ε4, PRS-*APOE*, and LIBRA2 with incident cases of dementia and AD, Cox models were applied to data of 345,785 UK Biobank participants at baseline (median follow-up: 13.8 years). Models were adjusted for age, sex, and other key covariates. Linear regression was used to examine associations with cognition (N=42,120) and dementia-related MRI markers (N = 31,180, such as fractional anisotropy/mean diffusivity, white matter hyperintensities, gray matter volume, hippocampus, and entorhinal cortex) among individuals without dementia.

**Results:** During follow-up, 6,300 participants developed dementia (132 cases per 100,000 person-years), including 2,803 cases of Alzheimer’s disease. All three predictors were independently associated with incident dementia and AD. Compared to non-carriers, *APOE* ε4 carriers had a threefold increased risk of dementia (HR = 3.02, 95% CI: 2.88–3.18) and over a fourfold increased risk of AD (HR = 4.33, 95% CI: 4.01–4.68). Each 1 standard deviation (SD) increase in PRS-APOE was associated with a 26% higher risk of dementia (HR = 1.26, 95% CI: 1.23–1.29) and a 35% higher risk of AD (HR = 1.35, 95% CI: 1.30–1.40). Each 1-SD increase in LIBRA2 score was linked to a 33% increase in dementia risk (HR = 1.33, 95% CI: 1.30–1.36) and 20% for AD (HR = 1.20, 95% CI: 1.15–1.24). Beyond individual effects, we evaluated model-level discrimination using the C-index. Representative results for dementia showed modest improvement over the base model (C-index = 0.811) with PRS-APOE (0.816) or LIBRA2 (0.819), and greater improvement with *APOE* ε4 (0.836). Combining *APOE* ε4 and LIBRA2 yielded near-optimal discrimination (0.843), close to the full model (0.846). While *APOE* ε4 was the dominant predictor of dementia diagnosis, LIBRA2 showed unique strongest predictive power for processing speed and neuroimaging markers. LIBRA2 was linked to a wide range of neuroimaging markers (e.g., WMH, WM integrity, grey matter, ventricles), while PRS-APOE was only associated with ventricular volume. *APOE* ε4 was related to couple of white matter measures. Significant LIBRA2-APOE interactions were observed in relation to both all-cause dementia and AD. The association between LIBRA2 and dementia risk was stronger among APOE ε4 non-carriers (interaction HR = 0.845, 95% CI: 0.807–0.884 for all-cause dementia; HR = 0.851, 95% CI: 0.792–0.914 for AD; both *p* adj < 0.001), suggesting that unfavorable lifestyle may have a greater impact in individuals without genetic risk. No significant interactions were found in cognitive or imaging models.

**Conclusion:** The *APOE* ε4 genotype was the strongest single predictor of dementia and cognitive outcomes. In contrast, the LIBRA2 index was most strongly associated with processing speed and neuroimaging markers, demonstrating its critical role in non-diagnostic brain health outcomes. These findings highlight the complementary and domain-specific contributions of genetic and lifestyle factors in dementia risk modeling.

## Introduction

Dementia is a clinical syndrome characterized by progressive cognitive decline that interferes with daily functioning and imposes a substantial socioeconomic burden worldwide (1).Among its various causes, Alzheimer’s disease (AD) is the most common form, accounting for approximately 60–80% of dementia cases. AD is marked by a long preclinical phase—often beginning 10 to 20 years before symptom onset—characterized by progressive neurodegenerative changes, including the hallmark pathological features of amyloid-beta plaques and tau tangles, and is frequently accompanied by vascular pathology such as white matter hyperintensities (WMH) (2–4). While age remains the strongest risk factor for dementia and AD, a growing body of research highlights the important roles of genetic susceptibility and modifiable lifestyle factors in influencing disease onset, cognitive health, and brain structure (5, 6). Among genetic factors, the apolipoprotein E (*APOE*) ε4 allele is the most well-established risk variant (particularly for the late onset form of AD), significantly increasing the risk of developing AD and impacting cognitive performance and neuroimaging markers such as white matter integrity and brain volume (7).

Beyond *APOE*, genome-wide association studies (GWAS) have identified numerous genetic loci associated with AD risk, enabling the construction of polygenic risk scores (PRSs) that estimate an individual’s cumulative genetic burden (8). PRSs are particularly promising for predicting disease risk, as they can be assessed early and at low cost, making them potential tools for identifying high-risk individuals who might benefit from targeted prevention (9, 10) and for enriching clinical trials with individuals at higher genetic risk (11). PRS derived from AD GWAS (AD-PRS)—excluding the *APOE* region—has demonstrated predictive value for AD beyond the *APOE* ε4 effect. AD-PRS has also been linked to neuroimaging markers of neurodegeneration and amyloid and tau pathology (12, 13).

Dementia risk is also shaped by lifestyle and health-related factors: approximately 45% of dementia burden is attributable to modifiable risk factors, many of which have a cardiometabolic or cardiovascular origin (including hypertension, physical inactivity, obesity, smoking, and diabetes) (14). A promising approach for assessing modifiable dementia risk is the LIBRA (“LIfestyle for BRAin Health”) index, which was developed in 2013 through a systematic review and Delphi expert study (15, 16). The original LIBRA index incorporated nine modifiable risk factors (coronary heart disease, physical inactivity, chronic kidney disease, diabetes, hypercholesterolemia, smoking, midlife obesity, midlife hypertension, and depression) and three protective factors (low-to-moderate alcohol intake, healthy diet, and high cognitive activity) for dementia. This index was recently updated to include additional risk factors (hearing impairment, little social contact, and sleep disturbances), resulting in the LIBRA2 index (17, 18). A higher (worse) LIBRA2 score has been consistently associated with an increased risk of cognitive decline, incident cognitive impairment, and dementia (16, 18–25). Furthermore, higher LIBRA scores have been linked to a greater burden of white matter hyperintensities (WMH), underscoring the potential connection between lifestyle factors and cerebrovascular health(26).

While both polygenic and lifestyle-based risk scores have demonstrated independent predictive value (22, 27), evidence regarding their combined utility remains mixed. Interestingly, studies in other disease areas have yielded divergent findings. For instance, in coronary heart disease, the addition of a PRS to a clinical risk model did not meaningfully improve risk prediction among white middle-aged adults (28). Conversely, in type 2 diabetes, although clinical risk scores generally outperformed PRS, adding PRS to clinical models significantly improved risk classification performance (29). These findings suggest that the relative contribution of genetic and lifestyle factors to disease risk—and their potential for integration—may differ across diseases. These inconsistencies across disease areas make it difficult to assume that genetic and lifestyle factors would interact or combine similarly in dementia. Moreover, dementia—particularly Alzheimer’s disease—has a more complex etiology than many other chronic conditions, involving hallmark neurodegenerative changes, and in many cases, also exhibits cerebrovascular pathology. Whether combining modifiable lifestyle factors with genetic risk enhances predictive accuracy for dementia remains unclear. Yet, insight into this is clinically relevant because identifying individuals at highest risk—especially those with both genetic susceptibility and unfavorable lifestyle profiles—could help guide earlier and more personalized prevention efforts. Therefore, to address this knowledge gap, we systematically compared the predictive value of *APOE* ε4 status, AD-PRS (excluding the *APOE* region), and the LIBRA2 lifestyle index—both individually and in combination— across three key outcome domains: incident dementia (including AD), cognitive performance, and neuroimaging markers.

Thus, we aimed to comprehensively compare the predictive value of AD-PRS (excluding the *APOE* region), *APOE* status, and the LIBRA2 index for incident dementia, as well as to examine their associations with cognitive performance and neuroimaging markers. Our study therefore integrates these genetic and lifestyle measures to clarify their relative importance for dementia and related phenotypes, providing a stronger evidence base for personalized prevention.

## Methods

### Participants

The current study utilizes data from the UK Biobank (UKB), which is a large, community-based, prospective cohort study (30). Briefly, UKB includes more than 500,000 participants aged 40-69 recruited between 2006 and 2010 and comprises 22 different assessment centers across the United Kingdom. The current analyses were conducted under UKB application 55392 using data from the July 2023 release.

### Measures

#### Incident dementia and Alzheimer’s disease (AD) ascertainment

Incident cases of all-cause dementia and AD were identified using algorithmically defined outcomes (ADOs) developed by the UKB in collaboration with clinical experts (31). These ADOs apply standardized methods to identify the earliest recorded cases of all-cause dementia and Alzheimer’s disease (AD) using three primary data sources: (1) hospital inpatient records with ICD-9/ICD-10 codes, (2) death register data, and (3) self-reported diagnoses from baseline assessment (used only to exclude prevalent cases). The event date was defined as the earliest recorded diagnosis of dementia or AD, while censoring occurred at the earliest of the date of death, date lost to follow-up, or end of follow-up (December 31, 2022). Time-to-event variables were calculated accordingly, and binary event indicators were created for dementia and AD. This approach follows UKB protocols and has demonstrated acceptable validity, with reported positive predictive values ranging from 57–100% for AD and around 67% for all-cause dementia, supporting its suitability for epidemiological research (32).

#### Cognitive assessments

Nine cognitive tests (see **Supplementary Table 1** for details) were selected from the UKB imaging visit (instance 2) to compute domain-specific and overall cognition scores, following similar methodologies to a previous study (33). Raw scores for each cognitive task were first examined for skewness; positively skewed distributions were log-transformed as appropriate. For tasks where lower scores indicated better performance, scores were reverse-coded (multiplied by −1) to ensure that higher values consistently reflected better cognitive performance. Subsequently, z-scores were calculated for each task. Values of each cognitive test exceeding ±6 standard deviations were winsorized (i.e., set to the nearest value within this range) to mitigate the influence of extreme outliers.

Composite scores for memory, executive function, and processing speed/attention were derived by averaging the available standardized z-scores across tasks within each domain. An overall cognition score was then calculated as the average of the three domain scores, provided that at least two domains were non-missing.

#### Neuroimaging data acquisition and processing

Neuroimaging data were obtained from a subsample of participants who attended a follow-up visit that included brain MRI scanning (34). Imaging was conducted on 3T Siemens Skyra scanners across three MRI centers following a publicly available protocol [https://www.fmrib.ox.ac.uk/ukbiobank/protocol/V4_23092014.pdf], and images processed using a publicly available processing pipeline [https://biobank.ctsu.ox.ac.uk/crystal/crystal/docs/brain_mri.pdf]. Details regarding imaging-derived phenotypes and processing are described elsewhere (35).

We focused on a set of MRI markers known to be relevant for dementia and brain health. From the diffusion MRI data, we included global mean fractional anisotropy (FA) and mean diffusivity (MD), processed using the Tract-Based Spatial Statistics (TBSS) pipeline in the FMRIB Software Library (FSL) [https://biobank.ctsu.ox.ac.uk/crystal/crystal/docs/brain_mri.pdf]. White matter tracts were defined using 48 standard-space tract masks developed by Mori et al. at Johns Hopkins University (36). Structural MRI outcomes, derived from T1-weighted images, included gray matter (GM) volume, and several subcortical volumes: entorhinal cortex, hippocampus, and bilateral lateral ventricles (left and right). WMH volume was calculated using T1 and T2-FLAIR images and segmented with the Brain Intensity Abnormality Classification Algorithm (BIANCA) (37), following the approach described by Miller et al. (38).

All MRI variables were standardized (z-scored) for analysis to facilitate comparison across measures. Because of its right-skewed distribution, WMH volume was log-transformed before standardization.

#### LIBRA2 index

Weighted LIBRA2 scores were calculated using 14 of the 15 modifiable risk and protective factors for dementia, excluding high cognitive activity due to unavailable data in the UKB. The LIBRA2 score reflects a weighted sum of these factors, with weights derived from previously published meta-analyses (15). Higher scores (range: –6.1 to 25.7) represent a less favorable lifestyle profile and indicate a higher lifestyle-related risk of dementia. Each factor was dichotomized based on predefined cut-offs (18), and a detailed overview of the UKB variables used for operationalizing the 14 components is provided in **Supplementary Table 2**.

#### Genotyping, Polygenic Risk Score Calculation (Excluding *APOE*), and *APOE* ε4 Status

Detailed information about the UKB genotyping procedures are reported elsewhere (39). Briefly, 488,377 UKB participants were genotyped for 805,426 markers using either the UK BiLEVE Axiom Array (49,950 participants) or the UK Biobank Axiom Array (438,427 participants). We used version 3 of the imputed UKB dataset, which was imputed against the Haplotype Reference Consortium (HRC) reference panel. The BGEN file obtained from UKB [https://biobank.ctsu.ox.ac.uk/ukb/ukb/docs/bgen12formats.pdf] was converted to PLINK binary format for further analyses. Quality control filtering excluded SNPs with >10% missingness and those failing the Hardy–Weinberg equilibrium test at a threshold of p < 1 × 10^−9^. PRS for AD were generated using PRS-CS, a Bayesian regression framework that employs continuous shrinkage (CS) priors to infer posterior SNP effect sizes from GWAS summary statistics and an external linkage disequilibrium (LD) reference panel (40). SNPs located within ±1 Mb of the *APOE* gene (defined from 1 Mb upstream of *APOE* gene to 1 Mb downstream of *APOE* gene) were excluded prior to PRS calculation, from here on out labeled PRS_-*APOE*_. The *APOE* genotype was directly derived from the SNPs rs429358 and rs7412 (41). Based on these, ε4 allele carrier status was defined (carriers: individuals with at least one ε4 allele; non-carriers: individuals without ε4 alleles) and included as a categorical variable in subsequent analyses.

#### Statistical analyses

Participants with a diagnosis of dementia at baseline, incomplete data on the 14 LIBRA2 components, or third-degree or closer genetic relatedness (identified using the UKB’s ‘GreedyRelatedExclude’ algorithm) were excluded. The final analytical samples included only participants with complete data on all 14 LIBRA2 components, genetic data, relevant covariates (age, sex, Townsend deprivation index, and education), and no history of dementia at baseline. All analyses were conducted using a complete case approach, i.e., participants with missing data on any of the model-specific variables were excluded. We evaluated the association of LIBRA2 score, AD PRS_-*APOE*_, and *APOE* status across three outcome domains: (1) Predictive ability for incident dementia and AD; (2) associations with cognitive performance; and (3) associations with AD-related neuroimaging markers. Separate analytical samples were used for each of the three outcome domains, based on the availability of complete data. These samples differed in size, as the cognitive assessments and neuroimaging scans were conducted in smaller subsets of the UK Biobank cohort during later follow-up visits.

Cox proportional-hazards models were utilized to assess the associations of LIBRA2, PRS_-*APOE*_, and *APOE* ε4 status with incident dementia and AD. The linearity assumption for continuous predictors (LIBRA2 and PRS-APOE) was assessed using penalized splines in Cox models. No substantial deviations from linearity were observed; therefore, these variables were modeled as linear terms. Hazard ratios (HRs) and 95% confidence intervals (CIs) were reported. Linear regression models were applied to examine the associations of LIBRA2, PRS_-*APOE*_, and *APOE* ε4 status with cognitive outcomes and neuroimaging biomarkers among participants free of dementia. All continuous predictors were standardized (z-scored) to enable direct comparison of effect sizes across predictors.

To assess the unique and incremental contribution of each factor, we fitted a series of models. For both the Cox and linear regressions, these included: 1) univariable models with a single predictor, 2) pairwise models combining two predictors, and 3) a full multivariable model with all three predictors (LIBRA2, PRS-*APOE*, and *APOE* ε4 status). Model performance was evaluated using Harrell’s C-statistic (higher values indicate better discrimination) and the Akaike Information Criterion (AIC; lower values indicate better model fit) for Cox models, and adjusted R^2^ (higher values indicate greater explanatory power) and AIC for linear regression models. To account for multiple testing, Bonferroni correction was applied, setting a significance threshold of 0.0125 for cognitive outcomes (0.05/4) and 0.005 for neuroimaging biomarkers (0.05/10). To examine potential moderation effects, pairwise interaction terms were included in the models for all combinations of the main predictors: LIBRA2 × *APOE*4 status, LIBRA2 × PRS_-APOE_, and *APOE*4 status × PRS_-APOE_.

All models were adjusted for age, sex, education, Townsend deprivation index, assessment center, and the first 20 genetic principal components to account for population stratification (included in all models, including those without PRS, to ensure comparability). For neuroimaging models, volumetric outcomes were additionally adjusted for intracranial volume (ICV) to account for individual differences in head size. Cox models used the time of the baseline visit as the model’s origin and time-on-study as the timescale and were adjusted for age at baseline and were stratified by assessment center to allow for center-specific baseline hazard functions. Linear regression models included age at the time of imaging or cognitive assessment (instance 2) and the corresponding assessment center at that time point as covariates. All statistical analyses were performed using R version 4.4.1 (R Core Team, 2024).

Finally, sensitivity analyses were conducted to assess the robustness of our findings. First, analyses were repeated among participants of British White ancestry to reduce potential bias due to population stratification. Second, we recalculated LIBRA2 using unweighted component scores to evaluate the influence of score weighting. Third, for cognitive outcomes, analyses were restricted to participants with complete data across all cognitive domains to ensure comparability across domains. Fourth, to assess potential reverse causality, we repeated dementia analyses after excluding participants who developed dementia within the first 5 years of follow-up (and, in a more stringent analysis, within 10 years).

## Results

### Descriptive Statistics

After applying the exclusions mentioned above, the dementia prediction analysis (incl. AD) included 345,785 participants, the cognitive analysis included 42,120 participants, and the neuroimaging analysis included 31,180 participants. Descriptive statistics across the three analytical samples are provided in **Table 1**. The mean age ranged from 54.3 to 56.5 years, with approximately 47–49% female participants. A higher proportion of participants in the cognition and neuroimaging samples had a college/university degree (48%) compared to the dementia sample (35%). *APOE* ε4 carriers (1 or 2 alleles) accounted for around 28% across all samples. The distributions of LIBRA2 scores and AD PRS_-*APOE*_ were similar across groups. Most participants were of British White ethnicity (>95%).

**Table 1.**
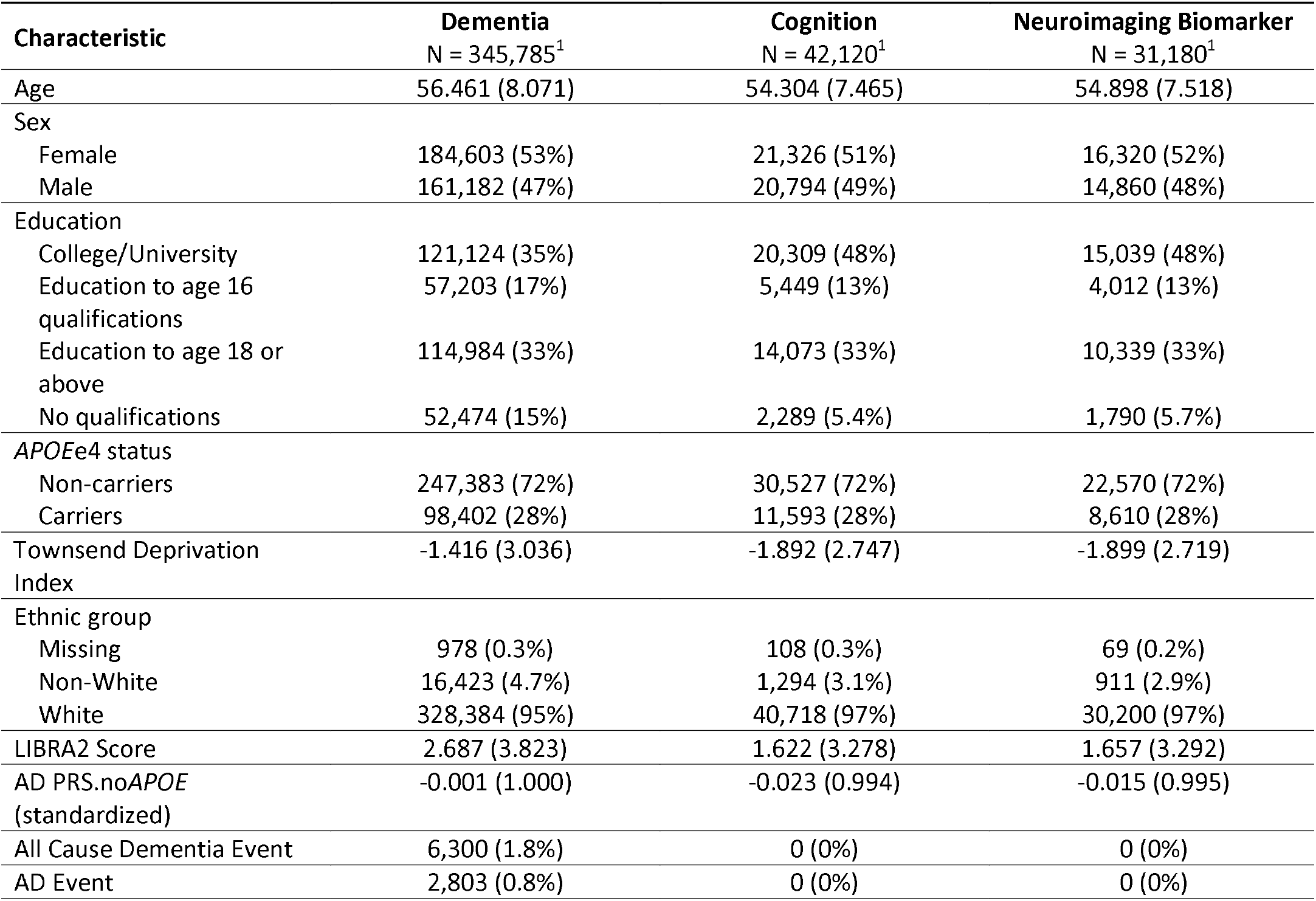
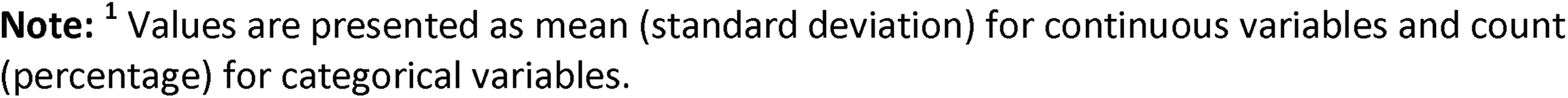
Baseline characteristics of UK Biobank participants included in the dementia, cognition, and neuroimaging biomarker analyses.

| Characteristic | Dementia<br>N = 345,785<sup>1</sup> | Cognition<br>N = 42,120<sup>1</sup> | Neuroimaging Biomarker<br>N = 31,180<sup>1</sup> |
| --- | --- | --- | --- |
| Age | 56.461 (8.071) | 54.304 (7.465) | 54.898 (7.518) |
| Sex |  |  |  |
| Female | 184,603 (53%) | 21,326 (51%) | 16,320 (52%) |
| Male | 161,182 (47%) | 20,794 (49%) | 14,860 (48%) |
| Education |  |  |  |
| College/University | 121,124 (35%) | 20,309 (48%) | 15,039 (48%) |
| Education to age 16<br>qualifications | 57,203 (17%) | 5,449 (13%) | 4,012 (13%) |
| Education to age 18 or<br>above | 114,984 (33%) | 14,073 (33%) | 10,339 (33%) |
| No qualifications | 52,474 (15%) | 2,289 (5.4%) | 1,790 (5.7%) |
| APOEε4 status | | | |
| Non-carriers | 247,383 (72%) | 30,527 (72%) | 22,570 (72%) |
| Carriers | 98,402 (28%) | 11,593 (28%) | 8,610 (28%) |
| Townsend Deprivation<br>Index | -1.416 (3.036) | -1.892 (2.747) | -1.899 (2.719) |
| Ethnic group |  |  |  |
| Missing | 978 (0.3%) | 108 (0.3%) | 69 (0.2%) |
| Non-White | 16,423 (4.7%) | 1,294 (3.1%) | 911 (2.9%) |
| White | 328,384 (95%) | 40,718 (97%) | 30,200 (97%) |
| LIBRA2 Score | 2.687 (3.823) | 1.622 (3.278) | 1.657 (3.292) |
| AD PRS.noAPOE<br>(standardized) | -0.001 (1.000) | -0.023 (0.994) | -0.015 (0.995) |
| All Cause Dementia Event | 6,300 (1.8%) | 0 (0%) | 0 (0%) |
| AD Event | 2,803 (0.8%) | 0 (0%) | 0 (0%) |

**Note:** <sup>1</sup> Values are presented as mean (standard deviation) for continuous variables and count (percentage) for categorical variables.

### *APOE* ε4 Status, AD PRS-*<SUB>APOE</SUB>*, and LIBRA2 in Relation to Dementia and Alzheimer’s Disease Risk

In Cox regression analyses, *APOE* ε4 status emerged as the strongest predictor of both all-cause dementia and AD. In multivariable Cox models, *APOE* ε4 carrier status (vs. non-carrier) was associated with a threefold increased risk of all-cause dementia (HR = 3.02, 95% CI: 2.88–3.18) and a more than fourfold increased risk of AD (HR = 4.33, 95% CI: 4.01–4.68) in multivariable models including all predictors. LIBRA2 score showed a stronger association with all-cause dementia, while AD PRS_-*APOE*_ demonstrated a more specific association with AD. each 1-SD increase in LIBRA2 score was associated with a 33% higher risk of all-cause dementia (HR = 1.33, 95% CI: 1.30–1.36), compared to a 20% increase in AD risk (HR = 1.20, 95% CI: 1.15–1.24). In contrast, each 1-SD increase in AD PRS was associated with a 26% higher risk of all-cause dementia (HR = 1.26, 95% CI: 1.23–1.29), and a relatively stronger 35% higher risk of AD (HR = 1.35, 95% CI: 1.30–1.40). All associations were highly significant (p < 0.001) (**Figure 1a**).

**Figure 1:**
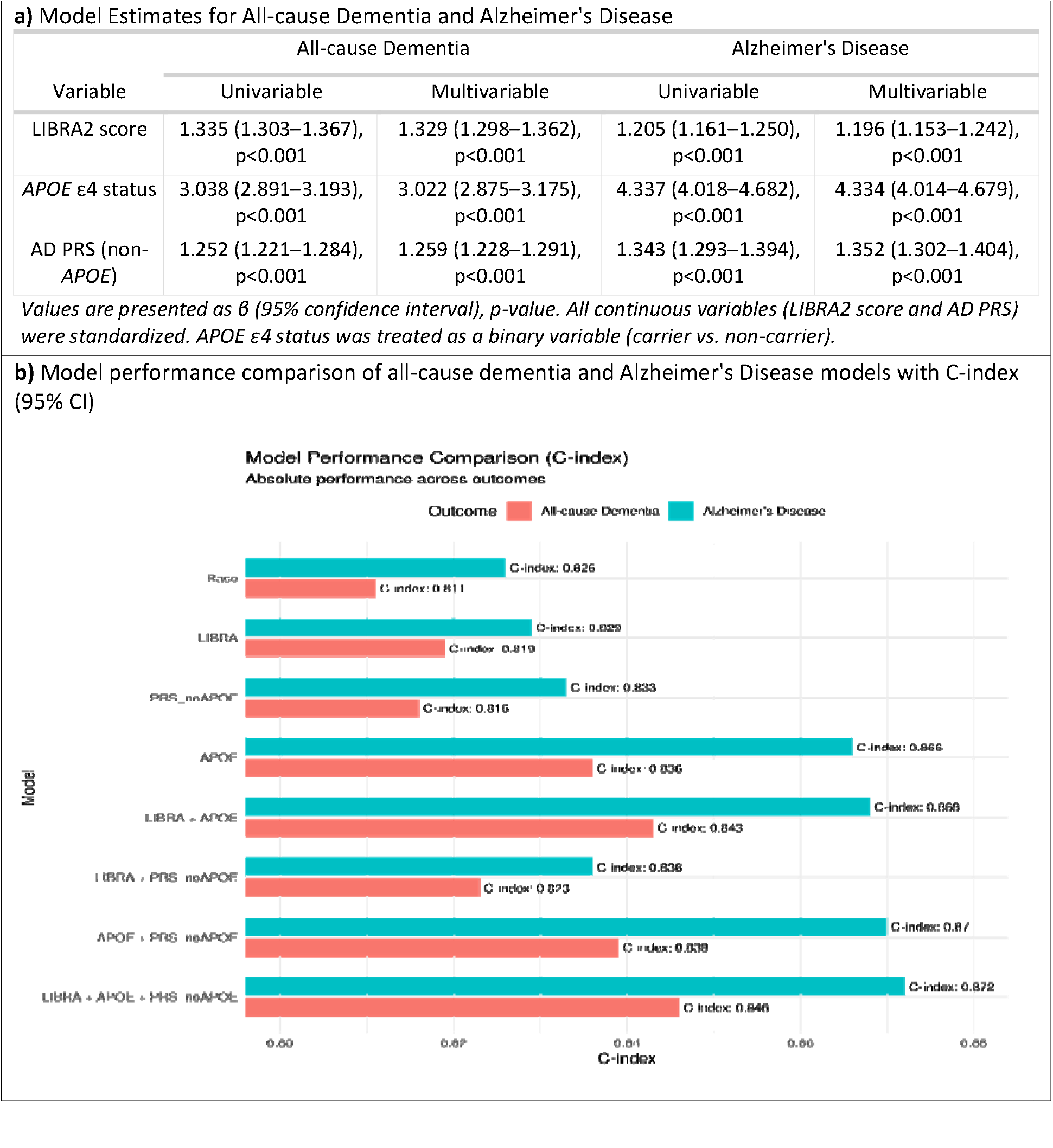
Cox Regression Model Estimates showing the Predictive Value of *APOE*, AD PRS, and LIBRA2 for All-cause Dementia and Alzheimer’s Disease. **Note:** Results are from Cox models adjusted for age, sex, education, deprivation index, assessment center (stratified), and 20 genetic principal components. △C: difference in the c-index (Harrell’s C-statistic) compared with the previous model; CI: confidence interval; LIBRA2: Lifestyle for Brain Health score; PRS-*APOE*: polygenic risk score for Alzheimer’s Disease excluding the *APOE* region

Eight models were compared for predicting all-cause dementia and AD using Harrell’s C-statistic. The base model (demographics + covariates) yielded a C-index of approximately 0.81 for dementia and 0.83 for AD. Adding individual predictors—LIBRA, PRS_-*APOE*_, or *APOE* ε4 status—incrementally improved model performance. Notably, *APOE* ε4 status yielded the largest individual improvement, increasing the C-index to 0.87 for AD and 0.85 for all-cause dementia. Combining predictors further improved performance. Among the two-predictor models, the combination of LIBRA2 and *APOE* ε4 status yielded the highest discriminative performance for all-cause dementia, with a C-index of 0.843, which was comparable to the full model including all three predictors (C-index = 0.846). For AD, three combination models—LIBRA2 + *APOE*, PRS_-*APOE*_ + *APOE*, and the full model including all three predictors— demonstrated similarly high predictive performance, with C-indices approaching 0.87. This suggests that *APOE* ε4 status, when combined with either LIBRA2 or PRS, captures most of the predictive power for AD (**Figure 1b**).

Interaction analyses revealed a statistically significant interaction between LIBRA2 score and *APOE* ε4 carrier status for both all-cause dementia (p_adj_ < 0.001) and AD (adjusted p-value, p_adj_ < 0.001). This suggests that the adverse effect of an unfavorable lifestyle on dementia risk was more pronounced among APOE ε4 non-carriers, while among carriers, the impact of lifestyle appeared attenuated, potentially due to a ceiling effect of high genetic risk.

A significant interaction was also observed between PRS_-*APOE*_ and *APOE* ε4 status for all-cause dementia (p_adj_ = 0.005), but not for AD after multiple testing correction (p_adj_ = 0.095). This suggests a synergistic effect of *APOE* and AD PRS on dementia risk (**Table 2**).

Kaplan–Meier survival curves (**Figure 2**) demonstrated clear and consistent separations in cumulative dementia-free survival for both all-cause dementia and AD based on *APOE* ε4 status, LIBRA2 score, PRS-*APOE*, and combined LIBRA2–*APOE* groups.

**Table 2.** Two-Way Interactions Between *APOE*, PRS, and LIBRA2 in Predicting Dementia Risk (Cox Models with Bonferroni-Adjusted p-values)

| Interaction Term | All-cause Dementia | Alzheimer's Disease |
| --- | --- | --- |
|  | HR (95% CI), p, p adj | HR (95% CI), p, p adj |
| LIBRA2: *APOE*_e4_status | 0.845 (0.807–0.884), p<0.001, p adj<0.001 | 0.851 (0.792–0.914), p<0.001, p adj<0.001 |
| LIBRA2: PRS-*APOE* | 0.974 (0.952–0.996), p=0.020, p adj=0.121 | 0.971 (0.937–1.005), p=0.096, p adj=0.574 |
| PRS-*APOE*: *APOE*_e4_status | 1.089 (1.036–1.144), p<0.001, p adj=0.005 | 1.099 (1.018–1.187), p=0.016, p adj=0.095 |

*Results are hazard ratios (HR) with 95% confidence intervals. All continuous variables were standardized prior to analysis. Bonferroni correction was applied for 6 comparisons.*

**Figure 2.**
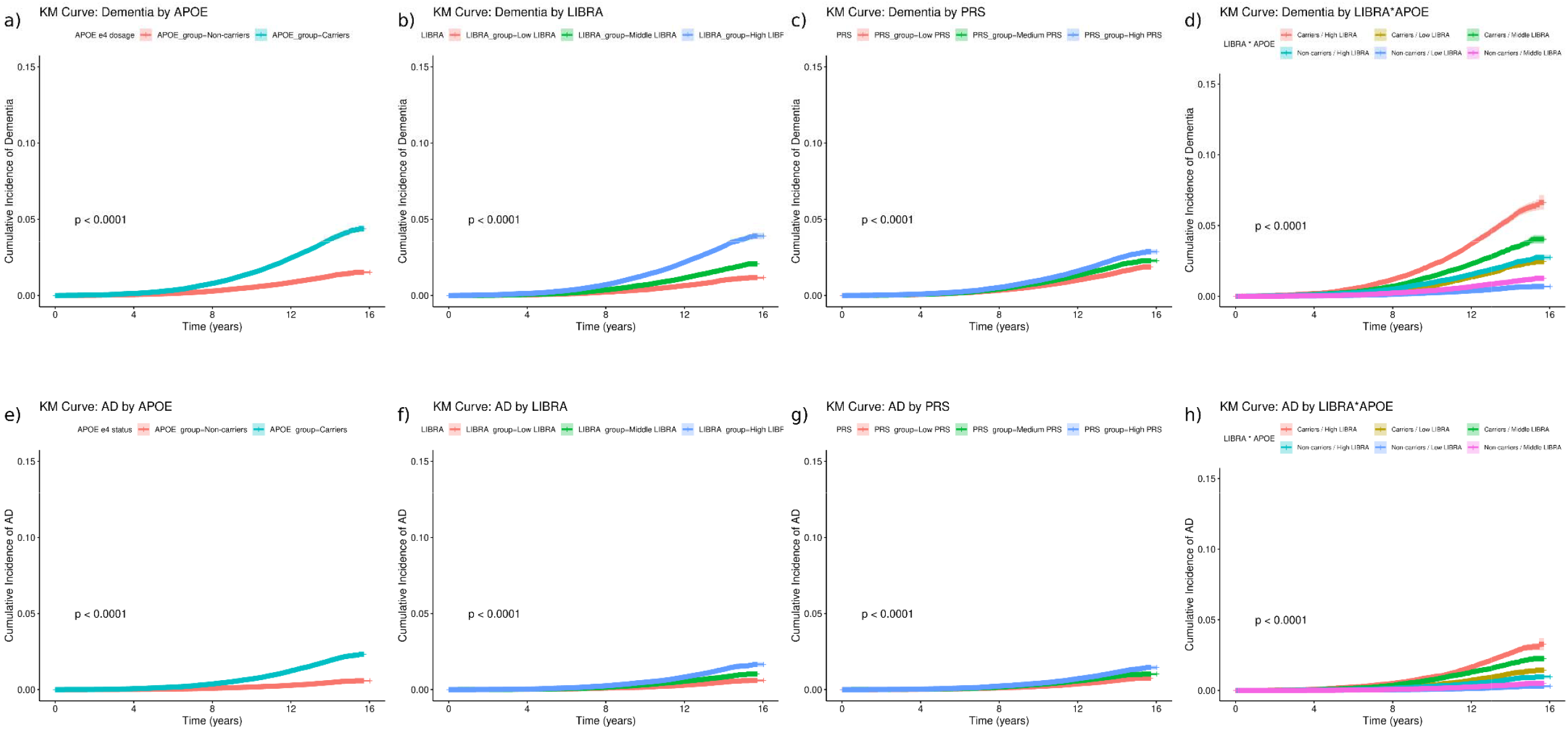
Kaplan–Meier (KM) curves showing the prediction of dementia (Panels A, B, C, and D) and Alzheimer’s disease (AD) (Panels E, F, G, and H) stratified by *APOE* status (carrier or non-carriers), LIBRA2 score (tertile 3), PRS (excluding the *APOE* region) (tertile 3), and by the combined LIBRA2 and *APOE* groups. *Abbreviations:* AD: Alzheimer’s disease; PRS: polygenic risk score excluding the *APOE* region; LIBRA2: Lifestyle for Brain Health score.

**Figure 3.**
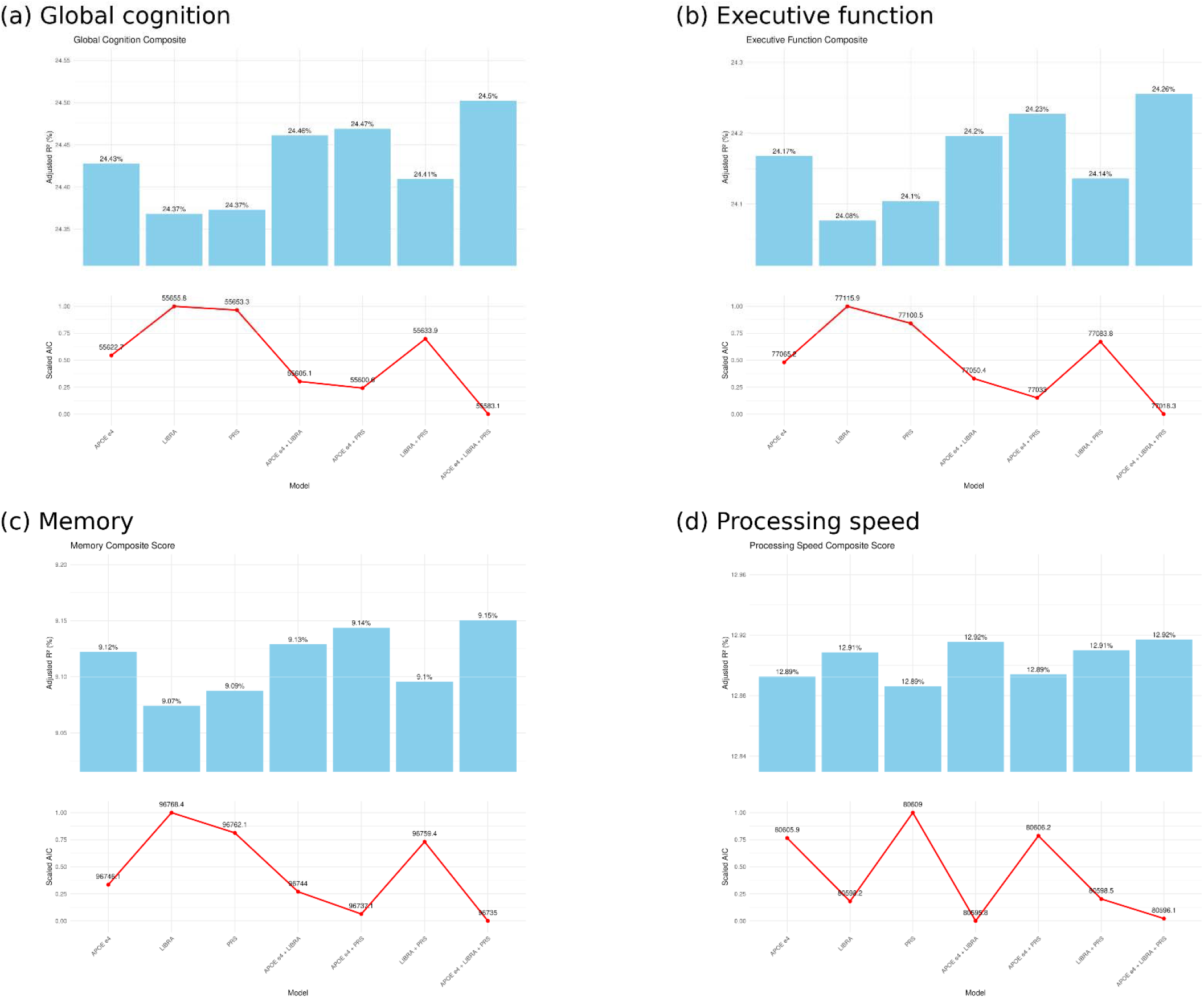
Comparison of Model Fit for Associations with Cognitive Domains. **Note**. *APOE*: Apolipoprotein E genotype; LIBRA2: Lifestyle for Brain Health score; PRS: polygenic risk score excluding the *APOE* region; CI: confidence interval; AIC: Akaike information criterion; p: uncorrected p-value; Bonferroni-corrected threshold for significance: p < 0.0125 (0.05/4); partial R^2^: proportion of variance explained by individual predictors; adjusted R^2^: variance explained by the whole model. In univariable models, *APOE* status and LIBRA2 score were nalyzed using their original units, while PRS was standardized (z-score) prior to analysis. In multivariable models, all predictors were standardized to facilitate effect size comparison.

### Associations of *APOE* ε4 Status, AD PRS_-*APOE*_, and LIBRA2 with Cognitive Function

Across all cognitive domains, *APOE* ε4 status consistently emerged as the strongest predictor of cognitive performance, explaining the largest proportion of variance (partial R^2^), except for processing speed, where LIBRA2 showed a stronger association (**Table 3**). In the multivariable models, *APOE* ε4 status had the largest effect sizes for overall cognition, executive function, and memory, with β values ranging from –0.054 to –0.037 (all p < 0.001). For processing speed, LIBRA2 showed the strongest effect, with a β (95% CI) of –0.011 (–0.017 to –0.005, p < 0.001). Similar trends were observed in the univariable models. No Bonferroni-corrected significant interactions were observed (**Supplementary Table 3**).

**Table 3.** Univariable and Multivariable Linear Regression Models for the Association of *APOE*, AD PRS, and LIBRA2 with cognitive performance.

|  | Univariable |  |  | Multivariable |  |  |
| --- | --- | --- | --- | --- | --- | --- |
| | $\beta$ (95% CI) | p | Partial $R^2$ | $\beta$ (95% CI) | p | Partial $R^2$ |
| **Overall cognition** |  |  |  |  |  |  |
| *APOE* ε4 status | -0.038 (-0.048, -0.028) | <0.001 | 0.130% | -0.037 (-0.047, -0.027) | <0.001 | 0.125% |
| LIBRA2 | -0.003 (-0.005, -0.002) | <0.001 | 0.051% | -0.011 (-0.015, -0.006) | <0.001 | 0.046% |
| PRS-*APOE* | -0.011 (-0.016, -0.007) | <0.001 | 0.057% | -0.011 (-0.016, -0.007) | <0.001 | 0.057% |
| **Memory** |  |  |  |  |  |  |
| *APOE* ε4 status | -0.043 (-0.060, -0.027) | <0.001 | 0.064% | -0.043 (-0.059, -0.027) | <0.001 | 0.063% |
| LIBRA2 | -0.003 (-0.005, -0.000) | 0.029 | 0.011% | -0.008 (-0.015, -0.000) | 0.042 | 0.010% |
| PRS-*APOE* | -0.013 (-0.020, -0.005) | <0.001 | 0.026% | -0.012 (-0.020, -0.005) | <0.001 | 0.026% |
| **Executive function** |  |  |  |  |  |  |
| *APOE* ε4 status | -0.055 (-0.068, -0.042) | <0.001 | 0.165% | -0.054 (-0.067, -0.041) | <0.001 | 0.160% |
| LIBRA2 | -0.004 (-0.006, -0.002) | <0.001 | 0.044% | -0.013 (-0.019, -0.007) | <0.001 | 0.040% |
| PRS-*APOE* | -0.017 (-0.023, -0.012) | <0.001 | 0.081% | -0.017 (-0.023, -0.012) | <0.001 | 0.081% |
| **Processing speed** |  |  |  |  |  |  |
| *APOE* ε4 status | -0.015 (-0.029, -0.002) | 0.028 | 0.011% | -0.014 (-0.028, -0.001) | 0.036 | 0.010% |
| LIBRA2 | -0.003 (-0.005, -0.002) | <0.001 | 0.030% | -0.011 (-0.017, -0.005) | <0.001 | 0.029% |
| PRS-*APOE* | -0.004 (-0.010, 0.002) | 0.190 | 0.004% | -0.004 (-0.010, 0.002) | 0.191 | 0.004% |

We constructed models that included LIBRA2, *APOE* ε4 status, and AD PRS-*APOE* both individually and in combination to examine their unique and joint contributions to cognitive performance. *APOE* ε4 status emerged as the strongest individual predictor for overall cognition, executive function, and memory. Among the two-predictor models, the combination of *APOE* and PRS-*APOE* yielded the highest performance, ranking second only to the full model, which includes all three predictors. In contrast, LIBRA2 remained the dominant predictor for processing speed, and the *APOE* + LIBRA2 model outperformed the other two-predictor combinations in this domain.

### Associations of *APOE* ε4 Status, AD PRS_-APOE_, and LIBRA2 with Neuroimaging Outcomes

We investigated the associations of *APOE* ε4 status, PRS, and the LIBRA2 with AD-related neuroimaging markers using univariable and multivariable linear regression models. Multivariable models adjusted for all predictors except that *APOE* was excluded from PRS models to avoid overlap (**Figure 4**).

**Figure 4.**
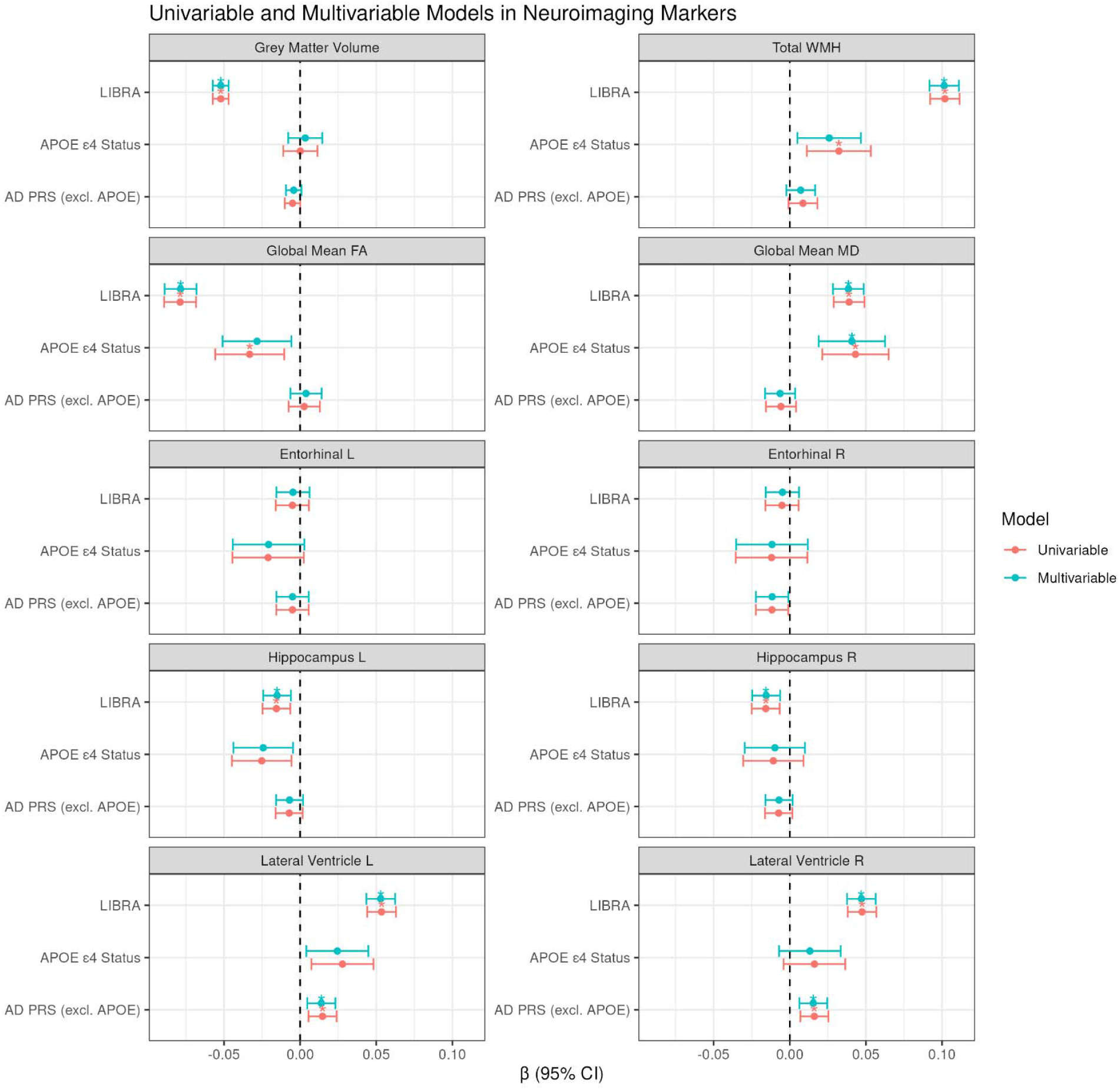
Associations between LIBRA2, genetic factors (PRS and *APOE*), and neuroimaging outcomes from univariable (red) and multivariable (green) linear regression models. **Note**. ^*^ Indicates significance after Bonferroni multiple testing correction. L: left hemisphere; R: right hemisphere; LIBRA2: Lifestyle for Brain Health score; PRS: polygenic risk score (excluding the *APOE* region); AD: Alzheimer’s disease; *APOE*: apolipoprotein E genotype; WMH: white matter hyperintensities; FA: fractional anisotropy; MD: mean diffusivity; CI: confidence interval. In univariable models, *APOE* status and LIBRA2 score were analyzed using their original units, while PRS was standardized (z-score). In multivariable models, all predictors were standardized to facilitate comparison of effect sizes.

In contrast to the findings for dementia incidence, LIBRA2 emerged as the strongest and most consistent predictor of neuroimaging markers.

Higher LIBRA2 scores were consistently associated with less favorable neuroimaging profiles. Specifically, LIBRA2 showed strong and significant associations with lower global FA, higher MD, greater WMH volume, and reduced grey matter and hippocampal volumes (all p < 0.001). These effects remained robust in multivariable models, indicating an independent influence of lifestyle on brain structure.

*APOE* ε4 status was significantly associated with lower FA, higher MD, and smaller hippocampal volume, but with lower effect sizes compared to LIBRA2. PRS showed no significant associations with most MRI outcomes, except for lateral ventricular volume, where both PRS (β = 0.014, p < 0.001) and LIBRA2 (β = 0.056, p < 0.001) demonstrated independent and strong associations. In contrast, *APOE* was not significant for this measure.

Overall, LIBRA2 emerged as the most consistent predictor across imaging domains. While *APOE* ε4 showed selective but modest effects, PRS contributed minimally, except for ventricular enlargement (details in **Supplementary Table 4**). No significant interactions (Bonferroni-adjusted) were observed for neuroimaging outcomes (**Supplementary Table 5**).

### Sensitivity analyses

Results from all sensitivity analyses were highly consistent with the main findings (Supplementary Tables 6–8). Specifically, restricting analyses to participants of British White ancestry, recalculating LIBRA2 using unweighted component scores, and— for dementia outcomes— excluding participants who developed dementia within the first 5 years of follow-up did not materially change the direction, magnitude, or relative ranking of the associations among APOE ε4 status, AD PRS, and LIBRA2. In all analyses, APOE ε4 remained the strongest predictor of dementia outcomes, whereas LIBRA2 remained the most consistent predictor of neuroimaging markers and of processing speed. No new significant interaction effects emerged in any sensitivity analysis.

## Discussion

This study evaluated the joint predictive value of APOE ε4, genome-wide AD polygenic scores (excluding APOE), and the updated LIBRA2 lifestyle index in relation to dementia, cognitive performance, and neuroimaging biomarkers in a general population. We found that APOE ε4 was the strongest predictor of dementia and cognitive outcomes, whereas LIBRA2 showed the most consistent associations with structural brain markers. Importantly, combining genetic and lifestyle factors modestly improved prediction beyond APOE alone, underscoring their distinct and complementary contributions. Notably, the observed interaction between LIBRA2 and APOE ε4 status in dementia prediction, but not in cognitive or imaging outcomes, suggests that gene–lifestyle interactions may be specific to dementia risk, with stronger associations between lifestyle and dementia observed in individuals without APOE ε4, but not evident for cognition or neuroimaging outcomes.

*APOE* ε4 carrier status emerged as the strongest individual predictor of both incident all-cause dementia and AD, with carriers showing over a threefold increased risk of all-cause dementia and more than a fourfold increased risk of AD— even after adjustment for PRS-*APOE* and LIBRA2. These results are consistent with previous studies showing that *APOE* ε4 status is more predictive of AD than broader genetic risk scores (13, 42, 43), reinforcing its central role in dementia risk stratification.

The PRS-*<SUB>APOE</SUB>*, constructed independently of the *APOE* region, showed moderate but significant associations with both dementia outcomes, with relatively stronger effects observed for AD than for all-cause dementia. Notably, we observed a significant interaction between PRS-*APOE* and *APOE* ε4 status for all-cause dementia, although the interaction for AD was not statistically significant after correction. This suggests a synergistic effect, where the contribution of polygenic risk may be amplified among *APOE* ε4 carriers. This gene–gene interaction is consistent with previous research indicating that genome-wide PRSs can improve AD risk stratification beyond *APOE* ε4 alone. While the standalone predictive utility of PRS remains lower than that of *APOE*, it may offer added value in refining risk estimates within genetically vulnerable subgroups (13). Biologically, there is emerging evidence that some GWAS-identified AD risk variants may influence *APOE* function through various mechanisms. These include: (1) linkage disequilibrium (LD) with *APOE* ε2/ε3/ε4 alleles, as seen in TOMM40 (44, 45) and APOC1; (2) regulatory variants such as rs405509, located in the *APOE* promoter region, which modulates *APOE* transcription (46, 47); and (3) variants that affect *APOE*-related biological pathways, such as lipid metabolism and amyloid processing. For example, APOC1 encodes apolipoprotein C-I and is not only in LD with ε4 but also identified as a potential eQTL influencing *APOE* expression (48). These findings support the notion that certain components of the PRS may exert their effects in part by modifying *APOE* function or expression. This may help explain why PRS and *APOE* ε4 appear to interact statistically and biologically, and why PRS may enhance risk stratification, particularly within *APOE* ε4 carriers.

The LIBRA2 score, representing a cumulative index of modifiable lifestyle risk factors, was also independently associated with both all-cause dementia and AD. Interestingly, LIBRA2 showed a relatively stronger association with all-cause dementia than with AD (HR = 1.33 vs. 1.20), supporting the notion that lifestyle risk may contribute to a broader range of neurodegenerative and cerebrovascular pathways. We further observed a significant interaction between LIBRA2 and APOE ε4 status in the Cox models for both dementia outcomes, suggesting that the association between lifestyle and dementia risk differs by genetic background. Specifically, the adverse impact of higher LIBRA2 scores was weaker among APOE ε4 carriers, potentially reflecting a ceiling effect or reduced susceptibility to lifestyle variation in individuals with high genetic risk. These findings underscore the importance of using longitudinal models to detect gene–lifestyle interplay relevant to dementia outcomes. While previous studies have investigated potential interactions between genetic risk and modifiable lifestyle factors, the results have been mixed. For example, our study consistent with Lourida et al.(5), which found no evidence of interaction between polygenic risk and lifestyle in predicting dementia risk, suggesting that a healthy lifestyle is beneficial regardless of genetic background. However, Euffer et al. (22) reported associations between LIBRA and dementia across levels of both APOE and polygenic risk score, with no significant interaction, which contrasts with our observation of a significant LIBRA2 × APOE ε4 interaction. In contrast, Deckers et al. (19) observed a significant interaction between APOE ε4 status and late-life LIBRA scores, with stronger associations among non-carriers. Furthermore, Licher et al. identified interaction effects between polygenic risk and modifiable factors, where favorable profiles had attenuated protective effects in those at high genetic risk. Against this background, our findings provide further evidence that lifestyle–genetic interactions may be most pronounced in predicting incident dementia, particularly when using longitudinal designs. Notably, the interaction between LIBRA2 and APOE ε4 status was specific to dementia outcomes and not observed for intermediate phenotypes like brain structure or cognition. This discrepancy may reflect several factors. First, the cross-sectional design of the cognitive and neuroimaging analyses— where lifestyle was assessed at baseline and outcomes were measured only once at a later time point—limits the ability to capture within-person changes and temporal dynamics. Without longitudinal follow-up of intermediate outcomes, it is more difficult to detect cumulative or time-dependent effects of lifestyle, especially when they may unfold gradually over decades. Second, the MRI and cognitive subsamples are typically smaller, healthier, and younger than the full cohort used for dementia analyses, due to selection biases and MRI contraindications. This may lead to restricted variance in exposures and outcomes, reducing statistical power and potentially underestimating associations or interaction effects. Third, intermediate phenotypes like brain structure or cognitive test scores may be more susceptible to measurement noise, day-to-day variability, and confounding by transient health or behavioral factors. In contrast, clinical endpoints like incident dementia reflect the long-term accumulation of multiple pathophysiological processes, potentially making them more sensitive to gene–environment interplay. Together, these factors highlight the value of large-scale, longitudinal cohort designs for detecting subtle and complex interactions between genetic susceptibility and modifiable lifestyle factors over the life course.

Comparisons of model performance further confirmed these observations. *APOE* ε4 status alone produced the largest improvement in discriminatory power, especially for AD prediction. However, the combination of *APOE* and LIBRA2 outperformed the other two-predictor models for all-cause dementia and performed comparably to the full model that also included the AD PRS. This underscores the complementary predictive value of genetic and modifiable lifestyle factors, providing support for integrated risk models that include both immutable and actionable components. Our analysis of cognitive performance revealed that *APOE* ε4 status consistently emerged as the strongest predictor across most cognitive domains, including overall cognition, executive function, and memory. These findings align with prior research showing that *APOE* ε4 is not only a risk factor for dementia diagnosis but also associated with poorer cognitive performance in cognitively normal individuals. The effect sizes for *APOE* ε4 remained robust in multivariable models, indicating that its impact on cognition is largely independent of other genetic and lifestyle-related risk factors.

Notably, an exception to this pattern was observed in the processing speed domain, where the LIBRA score demonstrated the strongest association. This suggests that lifestyle factors may exert a greater influence on basic cognitive processing mechanisms, which are especially sensitive to cerebrovascular and metabolic health—both key pathways targeted by the LIBRA2 index. Indeed, processing speed has consistently been shown to decline in the presence of white matter hyperintensities and other manifestations of small vessel disease (49), and to be among the earliest and most affected cognitive domains in such contexts. These patterns are supported by converging evidence from neuroimaging and neuropsychological studies in aging and SVD populations (50). These findings support and extend previous results from The Maastricht Aging Study, which showed that higher baseline LIBRA scores predicted a steeper longitudinal decline in processing speed over time (16). Our results suggest that even after accounting for genetic predisposition, modifiable risk factors may significantly influence specific cognitive functions, which may have particular clinical relevance for early intervention.

Despite the strong main effects of *APOE* and LIBRA, no statistically significant interaction effects were found between *APOE* ε4 status and either LIBRA or PRS-*<SUB>APOE</SUB>* on cognitive performance. This indicates largely additive, rather than synergistic, effects of genetic and lifestyle factors on cognitive variability in mid- to late-life individuals without dementia. These findings emphasize the utility of addressing modifiable risk factors such as those captured by LIBRA— even in genetically vulnerable individuals—to potentially preserve cognitive function, particularly in domains sensitive to cerebrovascular and lifestyle influences like processing speed.

Our findings reveal that across a broad set of neuroimaging markers—including white matter integrity (FA, MD), grey matter volume, hippocampal volume, lateral ventricle size, and WMH—higher LIBRA2 scores were consistently associated with less favorable neuroimaging profiles across multiple markers—including white matter integrity (FA, MD), grey matter volume, hippocampal volume, lateral ventricle size, and WMH.. The strongest associations were observed with WMH and global GM volume, followed by white matter microstructure (FA and MD) and hippocampal volume, suggesting that LIBRA2 captures a spectrum of brain alterations spanning both white and grey matter regions. This extends previous findings from The Maastricht Study. Earlier analyses reported that higher LIBRA scores were associated with greater WMH burden in the general population and reduced grey matter volume in men only(26). More recently, DeJong et al. showed that brain-age gap and white matter connectivity partially mediated the relationship between lifestyle and cognition in the same cohort, with brain-age gap accounting for nearly 25% of the total association (51).Our results build on these studies by demonstrating that LIBRA2 is broadly associated with multiple neuroimaging markers across both sexes, including medial temporal structures and major white matter tracts. Importantly, these associations remained robust in fully adjusted models, even after accounting for *APOE* genotype and polygenic risk, suggesting that the impact of modifiable lifestyle factors on brain structure is largely independent of genetic vulnerability. The particularly strong link between LIBRA2 and white matter abnormalities aligns with its emphasis on cardiovascular and metabolic risk factors, which are known to disproportionately affect white matter integrity, especially through mechanisms related to cerebral small vessel disease (52, 53).

Genetic predictors, including *APOE* ε4 status and PRS, showed weaker and less consistent associations with neuroimaging traits. While *APOE* ε4 exhibited modest associations with hippocampal and white matter measures, AD PRS was generally not associated with structural outcomes, except in the case of lateral ventricular volume. Interestingly, several *APOE* and PRS genetic effects appeared more pronounced in the left hemisphere than the right, especially in hippocampal and entorhinal regions. This lateralized pattern may reflect asymmetrical vulnerability to genetic risk in AD-relevant regions, a phenomenon reported in prior studies but not yet fully understood (54, 55).

Most strikingly, both LIBRA2 and PRS demonstrated significant and independent associations with lateral ventricular enlargement, a hallmark of global brain atrophy. The absence of a corresponding *APOE* effect suggests that ventricular expansion may reflect a cumulative burden of diffuse neurodegeneration driven by both polygenic risk and sustained lifestyle exposures rather than *APOE* ε4 alone. This convergence of genetic and environmental influence on a global brain marker highlights the necessity of integrative risk models and positions lateral ventricular volume as a promising structural biomarker for capturing both genetic liability and modifiable vulnerability in preclinical stages of neurodegeneration. It should, however, be noted that lateral ventricular volume is a non-specific marker of brain health that is associated with neurodegenerative disorders, including but not limited to AD, vascular dementia / cerebral small vessel disease, schizophrenia, multiple sclerosis, and Parkinson’s Diseases (56–59); therefore, its predictive value for specific trajectories is limited.

Given that LIBRA2 showed the strongest associations with MRI markers, while *APOE* was more strongly linked to dementia and cognition, it is plausible that neuroimaging markers—particularly those reflecting white and grey matter integrity—serve as intermediaries linking modifiable lifestyle factors to cognitive decline. This aligns with prior research demonstrating that brain structure and connectivity mediate the relationship between lifestyle and cognition (51).

This study has several strengths. Firstly, it is the first large-scale investigation to jointly evaluate *APOE* ε4 status, AD PRS, and the updated LIBRA lifestyle index in predicting dementia and related phenotypes. Secondly, we used an updated version of the LIBRA index, which has been shown to have stronger predictive ability (18). Thirdly, the inclusion of multiple outcome domains—clinical diagnosis of dementia, cognitive performance, and neuroimaging biomarkers— allowed for a comprehensive assessment of genetic and lifestyle influences across different stages of disease progression. Fourthly, the large sample size and prospective design of the UK Biobank provided sufficient statistical power to detect subtle main and interaction effects, increasing the robustness of our findings. Nevertheless, a few study limitations should be noted. First, the UKB cohort is relatively healthy compared to the general population, which likely reduces the observed incidence of dementia and limits generalizability. Second, the calculation of the LIBRA score in this cohort did not include the cognitive activity component, which in the original LIBRA study had an intermediate weight; its absence may have slightly reduced the predictive accuracy of LIBRA2. Third, participants who underwent brain MRI were a further selected subgroup of the UKB cohort and tended to be healthier and more health-conscious, which might have introduced additional selection bias in the neuroimaging analyses. Fourth, our reliance on algorithmically-defined dementia outcomes—while pragmatic and scalable—may lead to case misclassification, potentially biasing effect estimates towards the null. Finally, although we applied Bonferroni correction to control for multiple testing, this approach is conservative, particularly given the strong correlations among cognitive and neuroimaging outcomes. While this strategy minimizes false positive findings, it may have reduced power to detect small effects, especially interaction effects. Therefore, null findings for interaction terms and some genetic associations with imaging markers should be interpreted cautiously rather than as definitive evidence for the absence of such effects. Future studies may consider alternative approaches, such as false discovery rate control or multivariate modeling.

Recent comparative evaluations have benchmarked dementia risk scores such as LIBRA, Cardiovascular Risk Factors, Aging and Incidence of Dementia (CAIDE), the Australian National University Alzheimer’s Disease Risk Index (ANU-ADRI), and the newer Cognitive Health and Dementia Risk Index (CogDrisk) tools, revealing comparable predictive performance in various cohorts (60). Building on these efforts, future research could examine whether the updated LIBRA2 index— featuring a broader range of modifiable factors—offers improved risk stratification or responsiveness to lifestyle interventions, particularly in younger or more diverse populations.

Including additional covariates like diet quality, social support, or more detailed measures of cognitive engagement could further refine lifestyle risk modeling. Moreover, analyses in younger or more diverse cohorts could provide insights into early or preclinical disease processes. Mediation analyses may be useful to evaluate whether neuroimaging markers or cognitive performance mediate the effects of genetic or lifestyle risk on dementia, providing insights into underlying pathways. Finally, it would be valuable to replicate these analyses in non-population-based or clinical cohorts to confirm generalizability across different settings.

In conclusion, in this large UKB cohort, *APOE* status consistently emerged as the strongest predictor of dementia and dementia-related phenotypes, including hippocampal atrophy. However, LIBRA demonstrated particular relevance for processing speed and for several MRI measures—particularly those capturing white matter integrity and WMH burden— highlighting the potential importance of modifiable lifestyle factors in maintaining brain health. Although combining genetic and lifestyle factors modestly improved predictive performance, the incremental gains were relatively small. Ultimately, our findings support a dual-pronged public health message: identifying individuals with high genetic risk while promoting brain-healthy lifestyles for all, given the distinct benefits of the latter for maintaining brain structure and specific cognitive functions.

## Supporting information

Suppemental main text

## Data Availability

All data produced in the present study are available upon reasonable request to the authors

## Acknowledgments

This research has been conducted using the UKB Resource under Application Number 55392. Author Y. Zhang was supported by the China Scholarship Council (CSC) from the Ministry of Education of P.R. China. Authors S. Guloksuz and B.P.F. Rutten received support from the YOUTH-GEMs project, funded by the European Union’s Horizon Europe program under the grant agreement number: 101057182. S. Guloksuz was supported by the Ophelia research project, ZonMw grant number: 636340001. D. van der Meer is supported by the Research Council of Norway (RCN #351751). We declare that there are no conflicts of interest among the authors.

