## Supplementary material for "Genetic versus Modifiable Risk: Predicting Dementia, Cognition, and Brain Structure in the UK Biobank": Suppemental main text

**Supplementary Materials for:**

**Comparing the Predictive Value of Genetic Predisposition and the Modifiable Dementia Risk Score for Dementia and Their Associations with Cognition and Neuroimaging Biomarkers**

### Supplementary Table 1. Cognitive tests included in each cognitive domain.

| **Cognitive domain** | **Name of test** | **Short description of measure used** | **UKB Field ID** | **Preprocessing steps** |
| --- | --- | --- | --- | --- |
| **Memory** | Paired associate learning | Number of word pairs correctly associated | (UKB field 20197.2.0) | z-score |
|  | Pairs matching | Number of incorrect matches entered | (UKB field 399.2.2) | log transformation, inverse scoring |
| **Processing speed / attention** | Reaction time | Mean time to identify correct matches over 12 rounds | (UKB field 20023.2.0) | log transformation, z-score, inverse scoring |
|  | Trails 1 | Time to complete the numeric path | (UKB field 6348.2.0) | log transformation, z-score, inverse scoring |
|  | Numeric memory | Maximum number of digits remembered correctly | (UKB field 4282.2.0) | z-score |
| **Executive function** | Matrix pattern completion | Number of matrix pattern block puzzles correctly solved | (UKB field 6373.2.0) | log, z-score |
|  | Symbol digit substitution | Number of symbol digit matches made correctly | (UKB field 23324.2.0) | z-score |
|  | Tower rearranging | Number of correct puzzles solved | (UKB field 21004.2.0) | z-score |
|  | Trails 2 minus trails 1 | Time to complete the alphanumeric path minus time to complete the numeric path (purer measure of executive function) | (UKB field 6350.2.0) | z-score, inverse scoring |

### Supplementary Table 2: UK Biobank data fields used for operationalizing LIBRA2 components.

| **LIBRA2 component** | **UK Biobank variables** | **RR/HR Dementia^a^** |
| --- | --- | --- |
| High alcohol intake | (1) Number of drinks: 20117, 1558, 1568, 1588, 1598, 1608 | 1.18 |
| Sleep disturbances | (1) Self-reported sleep disturbances: 1200 | 1.19 |
| Midlife hypertension | (1) Antihypertensive treatment: 6177, 6153  (2) Objective BP measurement: 4080, 4079, 93, 94  (3) ICD-10: 41270 [codes: I10, I15]  (4) Self-reported (doctor) diagnosis: 6150, 20002 [codes: 1065, 1072, 1073] | 1.20 |
| Healthy diet | (1) Healthy diet score: 1309, 1319, 1289, 1299, 1329, 1339, 1349, 1369, 1379, 1389, 1438, 1448, 1458, 1468 | 0.82 |
| Chronic kidney disease | (1) Objective GFR measurement: 30720, 31, 21003, 21000  (2) Objective ACR measurement: 30500, 30510 | 1.35 |
| High physical activity | (1) Time spent on physical activities: 884, 894, 904, 914 | 0.73 |
| Low social participation | (1) Social isolation score: 1031, 6160, 709 | 1.41 |
| Diabetes | (1) Antidiabetic treatment: 20003 [codes: 1140884600, 1140883066], 6177, 6153  (2) ICD-10: 41270 [codes: E109, E119, E129, E139, E149]  (3) Self-reported (doctor) diagnosis: 2443, 20002 [codes: 1220, 1221, 1222, 1223] | 1.43 |
| Midlife obesity | (1) Objective BMI measurement: 21001 | 1.45 |
| Hearing impairment | (1) Self-report hearing impairment: 2247 | 1.49 |
| Smoking | (1) Self-report smoking: 20116 | 1.52 |
| Cholesterol | (1) cholesterol-lowering treatment: 6177, 6153  (2) Objective cholesterol measurement 30690  (3) ICD-10: 41270 [code: E780]  (4) Self-reported diagnosis: 20002 [code: 1473] | 1.54 |
| Coronary heart disease | (1) ICD-10: 41270 [codes: I20, I21, I22, I24]  (2) Self-reported (doctor) diagnosis: 6150, 20002 [codes: I1075, I1076] | 1.55 |
| Depression | (1) PHQ-2: 2050, 2060  (2) ICD-10: 41270 [codes: F32, F33, F34, F38, F39]  (3) Self-reported diagnosis: 20002 [codes: 1286, 1531] | 1.98 |

Abbreviations: ACR, albumin-creatinine ratio; BMI, body mass index; BP, blood pressure; GFR, glomerular filtration rate; ICD-10, International Classification of Diseases, Tenth Revision; LIBRA2, updated LIfestyle for BRAin health; PHQ-2, Patient Health Questionnaire-2

^a^Risk estimates from meta-analyses used for the development of LIBRA2 [1].

### Supplementary Table 3: Interaction Effects of Genotype and Lifestyle on Cognitive Outcomes.

|  | global_cognition_composite | memory_composite_score | executive_function_composite | Processing_speed_composite_score |
| --- | --- | --- | --- | --- |
| APOE_e4_status × LIBRA2 | | | | |
| Beta (95% CI) | -0.002 (-0.012, 0.008) | -0.011 (-0.027, 0.006) | -0.003 (-0.016, 0.010) | 0.006 (-0.007, 0.020) |
| p-value | 0.630 | 0.205 | 0.643 | 0.365 |
| Partial_R_squared | 0.000 | 0.000 | 0.000 | 0.000 |
| R_squared | 0.245 | 0.092 | 0.243 | 0.130 |
| Adjusted_R_squared | 0.245 | 0.091 | 0.242 | 0.129 |
| AIC | 55606.9 | 96744.4 | 77052.2 | 80597.0 |
| PRS_total_z × LIBRA2 | | | | |
| Beta (95% CI) | -0.001 (-0.006, 0.003) | 0.002 (-0.005, 0.009) | -0.005 (-0.011, 0.001) | -0.001 (-0.007, 0.005) |
| p-value | 0.558 | 0.564 | 0.077 | 0.755 |
| Partial_R_squared | 0.000 | 0.000 | 0.000 | 0.000 |
| R_squared | 0.245 | 0.092 | 0.242 | 0.130 |
| Adjusted_R_squared | 0.244 | 0.091 | 0.241 | 0.129 |
| AIC | 55635.5 | 96761.1 | 77082.7 | 80600.4 |
| APOE_e4_status × PRS_total_z | | | | |
| Beta (95% CI) | -0.003 (-0.013, 0.007) | 0.001 (-0.016, 0.017) | -0.010 (-0.023, 0.003) | -0.000 (-0.014, 0.013) |
| p-value | 0.524 | 0.929 | 0.116 | 0.981 |
| Partial_R_squared | 0.000 | 0.000 | 0.000 | 0.000 |
| R_squared | 0.245 | 0.092 | 0.243 | 0.130 |
| Adjusted_R_squared | 0.245 | 0.091 | 0.242 | 0.129 |
| AIC | 55602.2 | 96739.1 | 77032.5 | 80608.2 |

**Note:** Results are from linear regression models including interaction terms, adjusted for relevant covariates.
β (95% CI) represents the regression coefficient and its 95% confidence interval for the interaction term. *p* is the p-value for the interaction term.
Partial R² indicates the proportion of variance explained by the interaction, calculated by comparing full and reduced models (with and without the interaction term).
R² and Adjusted R² represent the overall model fit. AIC denotes the Akaike Information Criterion for the full model.

### Supplementary Table 4: Univariable and Multivariable Models in neuroimaging markers.

|  | Univariable | | | | | Multivariable | | | | |
| --- | --- | --- | --- | --- | --- | --- | --- | --- | --- | --- |
|  | β (95% CI) | p | Partial R² | Adj. R² | AIC | β (95% CI) | p | Partial R² | Adj. R² | AIC |
| zscore_global_mean_fa | | | | | | | | | | |
| APOE_e4_dosage | -0.037 (-0.057, -0.017) | <0.001 | 0.041% | 17.183% | 82854.095 | -0.016 (-0.026, -0.006) | 0.002 | 0.032% | 17.761% | 82637.067 |
| LIBRA2 | -0.024 (-0.027, -0.021) | <0.001 | 0.712% | 17.739% | 82643.461 | -0.079 (-0.090, -0.069) | <0.001 | 0.704% | 17.761% | 82637.067 |
| PRS_total_z | 0.002 (-0.008, 0.012) | 0.706 | 0.000% | 17.149% | 82866.729 | 0.003 (-0.007, 0.013) | 0.537 | 0.001% | 17.761% | 82637.067 |
| zscore_global_mean_md | | | | | | | | | | |
| APOE_e4_dosage | 0.047 (0.028, 0.067) | <0.001 | 0.074% | 23.362% | 80429.986 | 0.023 (0.013, 0.033) | <0.001 | 0.068% | 23.504% | 80373.945 |
| LIBRA2 | 0.012 (0.009, 0.015) | <0.001 | 0.194% | 23.454% | 80392.393 | 0.040 (0.029, 0.050) | <0.001 | 0.189% | 23.504% | 80373.945 |
| PRS_total_z | -0.005 (-0.015, 0.005) | 0.333 | 0.003% | 23.307% | 80452.301 | -0.006 (-0.015, 0.004) | 0.270 | 0.004% | 23.504% | 80373.945 |
| zscore_log_total_wmh | | | | | | | | | | |
| APOE_e4_dosage | 0.037 (0.018, 0.056) | <0.001 | 0.049% | 28.874% | 78097.905 | 0.016 (0.006, 0.025) | <0.001 | 0.035% | 29.828% | 77677.621 |
| LIBRA2 | 0.031 (0.028, 0.034) | <0.001 | 1.353% | 29.802% | 77687.235 | 0.101 (0.092, 0.111) | <0.001 | 1.337% | 29.828% | 77677.621 |
| PRS_total_z | 0.009 (-0.000, 0.019) | 0.058 | 0.011% | 28.847% | 78109.489 | 0.008 (-0.002, 0.017) | 0.114 | 0.008% | 29.828% | 77677.621 |
| zscore_volume_of_entorhinal_left_hemisphere_f27208_2_0 | | | | | | | | | | |
| APOE_e4_dosage | -0.023 (-0.044, -0.002) | 0.029 | 0.015% | 11.103% | 85069.765 | -0.011 (-0.022, -0.001) | 0.032 | 0.015% | 11.103% | 85071.541 |
| LIBRA2 | -0.002 (-0.005, 0.002) | 0.316 | 0.003% | 11.092% | 85073.507 | -0.005 (-0.016, 0.006) | 0.355 | 0.003% | 11.103% | 85071.541 |
| PRS_total_z | -0.006 (-0.017, 0.004) | 0.240 | 0.004% | 11.093% | 85073.131 | -0.006 (-0.017, 0.004) | 0.248 | 0.004% | 11.103% | 85071.541 |
| zscore_volume_of_entorhinal_right_hemisphere_f27301_2_0 | | | | | | | | | | |
| APOE_e4_dosage | -0.013 (-0.034, 0.008) | 0.212 | 0.005% | 10.612% | 85241.726 | -0.007 (-0.017, 0.004) | 0.225 | 0.005% | 10.625% | 85239.331 |
| LIBRA2 | -0.002 (-0.005, 0.002) | 0.320 | 0.003% | 10.611% | 85242.295 | -0.005 (-0.016, 0.006) | 0.355 | 0.003% | 10.625% | 85239.331 |
| PRS_total_z | -0.013 (-0.023, -0.002) | 0.018 | 0.018% | 10.624% | 85237.727 | -0.013 (-0.023, -0.002) | 0.019 | 0.018% | 10.625% | 85239.331 |
| zscore_volume_of_grey_matter_f25006_2_0 | | | | | | | | | | |
| APOE_e4_dosage | 0.002 (-0.008, 0.012) | 0.628 | 0.001% | 79.503% | 39203.631 | 0.003 (-0.002, 0.008) | 0.289 | 0.004% | 79.761% | 38809.425 |
| LIBRA2 | -0.016 (-0.018, -0.014) | <0.001 | 1.252% | 79.759% | 38810.126 | -0.053 (-0.058, -0.047) | <0.001 | 1.251% | 79.761% | 38809.425 |
| PRS_total_z | -0.006 (-0.011, -0.001) | 0.029 | 0.015% | 79.506% | 39199.116 | -0.005 (-0.010, 0.000) | 0.058 | 0.011% | 79.761% | 38809.425 |
| zscore_volume_of_hippocampus_left_hemisphere_f26562_2_0 | | | | | | | | | | |
| APOE_e4_dosage | -0.035 (-0.053, -0.018) | <0.001 | 0.051% | 37.950% | 73830.381 | -0.017 (-0.026, -0.009) | <0.001 | 0.048% | 37.976% | 73819.307 |
| LIBRA2 | -0.005 (-0.008, -0.002) | <0.001 | 0.039% | 37.942% | 73834.160 | -0.015 (-0.025, -0.006) | <0.001 | 0.036% | 37.976% | 73819.307 |
| PRS_total_z | -0.009 (-0.018, -0.000) | 0.047 | 0.013% | 37.926% | 73842.312 | -0.009 (-0.017, 0.000) | 0.054 | 0.012% | 37.976% | 73819.307 |
| zscore_volume_of_hippocampus_right_hemisphere_f26593_2_0 | | | | | | | | | | |
| APOE_e4_dosage | -0.020 (-0.038, -0.003) | 0.022 | 0.017% | 36.690% | 74458.475 | -0.010 (-0.019, -0.001) | 0.029 | 0.015% | 36.718% | 74446.761 |
| LIBRA2 | -0.005 (-0.008, -0.002) | <0.001 | 0.040% | 36.705% | 74451.137 | -0.016 (-0.025, -0.007) | <0.001 | 0.038% | 36.718% | 74446.761 |
| PRS_total_z | -0.009 (-0.018, 0.000) | 0.051 | 0.012% | 36.687% | 74459.881 | -0.009 (-0.017, 0.000) | 0.058 | 0.011% | 36.718% | 74446.761 |
| zscore_volume_of_lateralventricle_left_hemisphere_f26554_2_0 | | | | | | | | | | |
| APOE_e4_dosage | 0.024 (0.006, 0.042) | 0.009 | 0.022% | 32.809% | 76318.381 | 0.011 (0.002, 0.020) | 0.021 | 0.017% | 33.092% | 76188.577 |
| LIBRA2 | 0.016 (0.014, 0.019) | <0.001 | 0.400% | 33.064% | 76199.873 | 0.053 (0.044, 0.063) | <0.001 | 0.392% | 33.092% | 76188.577 |
| PRS_total_z | 0.016 (0.006, 0.025) | <0.001 | 0.035% | 32.818% | 76314.253 | 0.015 (0.006, 0.024) | 0.002 | 0.032% | 33.092% | 76188.577 |
| zscore_volume_of_lateralventricle_right_hemisphere_f26585_2_0 | | | | | | | | | | |
| APOE_e4_dosage | 0.015 (-0.003, 0.033) | 0.104 | 0.008% | 33.626% | 75936.101 | 0.006 (-0.003, 0.015) | 0.183 | 0.006% | 33.858% | 75828.651 |
| LIBRA2 | 0.015 (0.012, 0.017) | <0.001 | 0.319% | 33.832% | 75838.727 | 0.047 (0.038, 0.057) | <0.001 | 0.313% | 33.858% | 75828.651 |
| PRS_total_z | 0.017 (0.008, 0.026) | <0.001 | 0.043% | 33.649% | 75925.399 | 0.016 (0.007, 0.025) | <0.001 | 0.039% | 33.858% | 75828.651 |

### Supplementary Table 5: Interaction Effects of Genotype and Lifestyle on Neuroimaging Markers.

|  | APOE4 * LIBRA2 | | | | | | AD PRS (excl. APOE) * LIBRA2 | | | | | | PRS (excl. APOE) * APOE | | | | | |
| --- | --- | --- | --- | --- | --- | --- | --- | --- | --- | --- | --- | --- | --- | --- | --- | --- | --- | --- |
|  | β (95% CI) | p | Partial R² | R² | Adj. R² | AIC | β (95% CI) | p | Partial R² | R² | Adj. R² | AIC | β (95% CI) | p | Partial R² | R² | Adj. R² | AIC |
| Entorhinal R | -0.000 (-0.024, 0.023) | 0.985 | 0.000% | 10.712% | 10.618% | 85021.155 | 0.008 (-0.002, 0.019) | 0.129 | 0.007% | 10.729% | 10.635% | 85015.223 | -0.012 (-0.036, 0.011) | 0.314 | 0.003% | 10.726% | 10.631% | 85016.310 |
| Entorhinal L | -0.001 (-0.024, 0.023) | 0.956 | 0.000% | 11.203% | 11.109% | 84849.174 | 0.004 (-0.006, 0.015) | 0.421 | 0.002% | 11.199% | 11.105% | 84850.686 | -0.001 (-0.024, 0.023) | 0.935 | 0.000% | 11.204% | 11.110% | 84849.052 |
| Hippocampus L | -0.004 (-0.023, 0.016) | 0.705 | 0.000% | 38.072% | 38.006% | 73612.681 | -0.000 (-0.009, 0.009) | 0.995 | 0.000% | 38.065% | 37.999% | 73616.344 | -0.010 (-0.030, 0.009) | 0.303 | 0.003% | 38.057% | 37.992% | 73620.072 |
| Hippocampus R | -0.002 (-0.021, 0.018) | 0.867 | 0.000% | 36.830% | 36.763% | 74231.758 | -0.001 (-0.010, 0.008) | 0.879 | 0.000% | 36.833% | 36.766% | 74230.310 | -0.011 (-0.030, 0.009) | 0.293 | 0.004% | 36.815% | 36.748% | 74239.287 |
| Global Mean FA | 0.005 (-0.018, 0.027) | 0.682 | 0.001% | 17.733% | 17.648% | 82465.739 | -0.005 (-0.015, 0.005) | 0.343 | 0.003% | 17.720% | 17.636% | 82470.509 | -0.012 (-0.035, 0.011) | 0.303 | 0.003% | 17.166% | 17.081% | 82679.729 |
| Global Mean MD | 0.009 (-0.013, 0.030) | 0.444 | 0.002% | 23.476% | 23.398% | 80209.173 | 0.000 (-0.009, 0.010) | 0.927 | 0.000% | 23.446% | 23.367% | 80221.609 | 0.016 (-0.006, 0.038) | 0.155 | 0.007% | 23.347% | 23.268% | 80261.916 |
| Grey Matter Volume | 0.005 (-0.006, 0.016) | 0.369 | 0.003% | 79.840% | 79.819% | 38619.408 | -0.001 (-0.006, 0.004) | 0.625 | 0.001% | 79.842% | 79.820% | 38617.630 | -0.003 (-0.014, 0.008) | 0.603 | 0.001% | 79.591% | 79.569% | 39003.313 |
| Total WMH | 0.007 (-0.013, 0.028) | 0.483 | 0.002% | 29.814% | 29.740% | 77515.435 | 0.001 (-0.008, 0.011) | 0.805 | 0.000% | 29.805% | 29.731% | 77519.564 | 0.008 (-0.013, 0.029) | 0.448 | 0.002% | 28.869% | 28.794% | 77932.496 |
| Lateral Ventricle L | -0.001 (-0.021, 0.019) | 0.922 | 0.000% | 33.074% | 33.003% | 76032.631 | 0.003 (-0.006, 0.012) | 0.514 | 0.001% | 33.082% | 33.011% | 76028.978 | 0.008 (-0.013, 0.028) | 0.463 | 0.002% | 32.835% | 32.764% | 76143.703 |
| Lateral Ventricle R | 0.003 (-0.017, 0.023) | 0.787 | 0.000% | 33.867% | 33.797% | 75660.945 | 0.007 (-0.002, 0.016) | 0.123 | 0.008% | 33.892% | 33.822% | 75649.358 | 0.014 (-0.006, 0.034) | 0.182 | 0.006% | 33.689% | 33.619% | 75744.639 |

**Note:** Linear regression models with interaction terms for APOE, AD PRS, and LIBRA2 were adjusted for relevant covariates. β (95% CI) and p refer to the interaction term. Partial R² was calculated by comparing models with and without the interaction. R², Adjusted R², and AIC reflect overall model fit.

### Supplementary Table 6: All-cause dementia and Alzheimer's Disease sensitivity (full Cox model)

|  | **Analysis** | **LIBRA2 (per SD) HR (95% CI), p** | **PRS-APOE (per SD) HR (95% CI), p** | **APOE ε4 HR (95% CI), p** | **LIBRA2 × APOE ε4 HR (95% CI), p** | **PRS × APOE ε4 HR (95% CI), p** | **LIBRA2 × PRS HR (95% CI), p** |
| --- | --- | --- | --- | --- | --- | --- | --- |
| **All-cause dementia** | Main | 1.33 (1.30–1.36), p<0.001 | 1.26 (1.23–1.29), p<0.001 | 3.02 (2.88–3.18), p<0.001 | 0.85 (0.81–0.88), p<0.001 | 1.09 (1.04–1.14), p<0.001 | 0.97 (0.95–1.00), p=0.020 |
|  | British White | 1.34 (1.31–1.37), p<0.001 | 1.27 (1.23–1.30), p<0.001 | 3.04 (2.90–3.20), p<0.001 | 0.84 (0.80–0.87), p<0.001 | 1.10 (1.05–1.15), p<0.001 | 0.98 (0.95–0.99), p=0.015 |
|  | Exclude first 5 years | 1.31 (1.25–1.33), p<0.001 | 1.23 (1.19–1.27), p=0.001 | 2.95 (2.78–3.13), p<0.001 | 0.86 (0.82–0.92), p<0.001 | 1.08 (1.02–1.14), p=0.009 | 0.98 (0.95–1.01), p=0.12 |
| **Alzheimer's Disease** | Main | 1.20 (1.15–1.24), p<0.001 | 1.35 (1.30–1.40), p<0.001 | 4.33 (4.01–4.68), p<0.001 | 0.85 (0.79–0.91), p<0.001 | 1.10 (1.02–1.19), p=0.016 | 0.97 (0.94–1.01), p=0.096 |
|  | British White | 1.21 (1.16–1.25), p<0.001 | 1.36 (1.30–1.41), p<0.001 | 4.32 (4.00–4.67), p<0.001 | 0.85 (0.79–0.91), p<0.001 | 1.11 (1.03–1.20), p=0.010 | 0.96 (0.93–1.00), p=0.085 |
|  | Exclude first 5 years | 1.18 (1.12–1.23), p<0.001 | 1.31 (1.25–1.37), p<0.001 | 4.10 (3.76–4.47), p<0.001 | 0.87 (0.80–0.95), p=0.003 | 1.08 (0.99–1.18), p=0.08 | 0.98 (0.94–1.02), p=0.22 |

### Supplementary Table 7: Cognition sensitivity （2 outcomes）

| **Outcome** | **Analysis** | **LIBRA2 β (95% CI), p** | **PRS-APOE β (95% CI), p** | **APOE ε4 β (95% CI), p** |
| --- | --- | --- | --- | --- |
| **Global cognition** | Main | −0.011 (−0.015 to −0.006), p<0.001 | −0.011 (−0.016 to −0.007), p<0.001 | −0.037 (−0.047 to −0.027), p<0.001 |
|  | British White | −0.011 (−0.014 to −0.006), p<0.001 | −0.011 (−0.016 to −0.007), p<0.001 | −0.036 (−0.047 to −0.026), p<0.001 |
|  | Unweighted LIBRA2 | −0.010 (−0.014 to −0.006), p<0.001 | −0.011 (−0.016 to −0.007), p<0.001 | −0.037 (−0.047 to −0.027), p<0.001 |
|  | Same-sample cognition | −0.012 (−0.017 to −0.007), p<0.001 | −0.012 (−0.016 to −0.008), p<0.001 | −0.039 (−0.049 to −0.029), p<0.001 |
| **Processing speed** | Main | −0.011 (−0.017 to −0.005), p<0.001 | −0.004 (−0.010 to 0.002), p=0.19 | −0.014 (−0.028 to −0.001), p=0.036 |
|  | British White | −0.011 (−0.017 to −0.005), p<0.001 | −0.005 (−0.010 to 0.002), p=0.12 | −0.015 (−0.029 to −0.001), p=0.031 |
|  | Unweighted LIBRA2 | −0.010 (−0.016 to −0.004), p=0.001 | −0.004 (−0.010 to 0.002), p=0.19 | −0.014 (−0.028 to −0.001), p=0.036 |
|  | Same-sample cognition | −0.012 (−0.018 to −0.005), p<0.001 | −0.004 (−0.010 to 0.002), p=0.19 | −0.016 (−0.030 to −0.002), p=0.028 |

### Supplementary Table 8: MRI sensitivity（3 outcomes）

| **Outcome** | **Analysis** | **LIBRA2 β (95% CI), p** | **PRS-APOE β (95% CI), p** | **APOE ε4 β (95% CI), p** |
| --- | --- | --- | --- | --- |
| **log(WMH+1)** | Main | 0.060 (0.052–0.068), p<0.001 | 0.004 (−0.003–0.011), p=0.26 | 0.010 (0.001–0.019), p=0.028 |
|  | British White | 0.059 (0.050–0.067), p<0.001 | 0.005 (−0.002–0.011), p=0.18 | 0.011 (0.002–0.019), p=0.022 |
|  | Unweighted LIBRA2 | 0.056 (0.048–0.064), p<0.001 | 0.004 (−0.003–0.011), p=0.26 | 0.010 (0.001–0.019), p=0.028 |
| **Hippocampus volume** | Main | −0.020 (−0.028 to −0.012), p<0.001 | −0.002 (−0.009–0.006), p=0.65 | −0.012 (−0.021 to −0.003), p=0.008 |
|  | British White | −0.020 (−0.028 to −0.012), p<0.001 | −0.002 (−0.009–0.006), p=0.72 | −0.011 (−0.021 to −0.002), p=0.012 |
|  | Unweighted LIBRA2 | −0.018 (−0.026 to −0.010), p<0.001 | −0.002 (−0.009–0.006), p=0.65 | −0.012 (−0.021 to −0.003), p=0.008 |
|  | Main | −0.035 (−0.043 to −0.027), p<0.001 | 0.002 (−0.005–0.009), p=0.58 | −0.010 (−0.019 to −0.001), p=0.030 |
| **Global FA** | British White | −0.034 (−0.042 to −0.026), p<0.001 | 0.003 (−0.004–0.010), p=0.45 | −0.010 (−0.019 to −0.001), p=0.048 |
|  | Unweighted LIBRA2 | −0.033 (−0.041 to −0.025), p<0.001 | 0.002 (−0.005–0.009), p=0.58 | −0.010 (−0.019 to −0.001), p=0.030 |
